# Estimating the size of the COVID-19 outbreak in Italy: Application of an exponential decay model to the weighted and cumulative average daily growth rate

**DOI:** 10.1101/2020.05.20.20108241

**Authors:** Nicola Bartolomeo, Paolo Trerotoli, Gabriella Serio

## Abstract

To estimate the size of the novel coronavirus (COVID-19) outbreak in Italy, this paper introduces the cumulated and weighted average daily growth rate (WR) to evaluate an epidemic curve, On the basis of an exponential decay model (EDM), we provide estimations of the WR in four-time intervals from February 27 to April 07, 2020. By calibrating the parameters of the EDM to the reported data in Hubei Province of China, we also attempt to forecast the evolution of the outbreak. We compare the EDM applied to WR and the Gompertz model, which is based on exponential decay and is often used to estimate cumulative events. Specifically, we assess the performance of each model to short-term forecast of the epidemic, and to predict the final epidemic size.

Based on the official counts for confirmed cases, the model applied to data from February 27 until the 17st of March estimate that the cumulative number of infected could reach 131,280 (with a credibility interval 71,415-263,501) by April 25 (credibility interval April 12 to May 3). With the data available until the 24st of March the peak date should be reached on May 3 (April 23 to May 23) with 197,179 cumulative infections expected (130033-315,269); with data available until the 31st of March the peak should be reached on May 4 (April 25 to May 18) with 202,210 cumulative infections expected (155.235-270,737); with data available until the 07st of April the peak should be reached on May 3 (April 26 to May 11) with 191,586 (160,861-232,023) cumulative infections expected. Based on the average mean absolute percentage error (MAPE), cumulated infections forecasts provided by the EDM applied to WR performed better across all scenarios than the Gompertz model.

An exponential decay model applied to the cumulated and weighted average daily growth rate appears to be useful in estimating the number of cases and peak of the COVID-19 outbreak in Italy and the model was more reliable in the exponential growth phase.

## Introduction

In December 2019, an outbreak of a novel coronavirus (COVID-19) occurred in Hubei Province, China. Since then, COVID-19 has spread around the world, and on March 11, 2020, it was declared a pandemic by the World Health Organization (WHO). As of April 30, 2020, there have been 3,090,445 cases and 217,769 deaths, reported globally. Italy has been one of the worst affected countries, and so far has reported 203,951 cases and 27682 deaths (as of April 30, 2020) [1]. The Italian government has implemented a series of social distancing and public health strategies to try and reduce the spread of COVID-19, including closing schools, screening passengers at railway stations and airports, identifying active cases, and isolating suspected cases. On March 9, 2020, Italy became the first country to implement a nation-wide lockdown to try and limit the COVID-19 spread [2].

While the transmission potential of COVID-19 can be high [3,4], the epidemiological features of COVID-19 are still unclear. Differences in the reporting of cases and deaths complicate any analysis of the COVID-19 epidemic. Governments need to be able to accurately estimate cases in a geographical area so that social distancing and public health measures can be implemented quickly. Besides, it is essential to formulate a plan to ensure that patients with COVID-19 can be cared for in the hospital if required.

Many scholars have been attempting to estimate the basic reproduction number (R0) and predict the future trajectory of the COVID-2019 outbreak. In this paper, we use phenomenological models without detailed microdata, which allows for simple calibrations to be made to the reported empirical data so that quick interpretations can be formulated.

Many authors have attempted to use the cumulative number of new infections to estimate the “peak” of the outbreak with the main models used in epidemiology, SIR, SEIR, Log-Linear, Negative-Binomial. However, these forecasting models are limited in that the transmission speed of new viruses, such as SARS-CoV-2 is unknown. As such, many epidemiologists have reported that it is challenging to make medium and long-term predictions regarding the peak.

Studying the spread of COVID-19 in a country such as China could be useful. In particular, Hubei Province, which is similar to Italy in terms of resident population numbers, could provide vital information for exploring the dynamics of COVID-19, despite the social contexts between the two locations being very different.

The contagion curve of China and/or the Hubei Province could be used to create epidemiological growth models, from which the parameters could be used as a starting point for other populations. However, the absolute number of infections depends on a number of population-based characteristics, such as ethnicity and age structure.

Another useful indicator to evaluate the speed of the virus spread, which can be used comparatively between two populations, is the daily growth rate and its associated trend. The daily growth rate, which is the number of new cases at time t divided by the total number of cases recorded up to time t-1, is a useful indicator for assessing the daily spread and speed of infection. The peak of an epidemic is considered to be when the growth rate is zero. This rate can be comparatively used as it is affected by the intrinsic characteristics of the virus. However, it may be affected by the particular detection conditions in a single day (number of tests carried out, time of detection, delay in the processing of tests by laboratories, partial communication of daily data by regions, and endogenous situations, such as climatic events that influence the movements of people). Indeed, the daily growth rate can have significant variability, particularly if the rate is only tracked over a short period of time. Besides, the daily growth rate and its variability depend on the total number of infections. At the beginning of the outbreak, the daily rates are more unstable, but they appear to stabilize when the cumulative number of infections increases. The excessive variability of daily growth rates adversely affects the robustness of forecasting models.

The average of the daily growth rate could be used to reduce this high variability in the daily growth rate, as it has a smoothing effect on the rates cited.

This study aimed to define and evaluate the daily growth rate and apply the cumulative and weighted average of the crude daily growth rate to an epidemic curve.

## Material and Methods

Italian epidemic data registered between February 26, 2020 and April 18, 2020 by Italian Protezione Civile [5] and Chinese (Hubei Province) epidemic data, between January 21, 2020 and March 18, 2020, were taken from Hubei Province government website and used in this study [6].

The first step was to evaluate the growth rate: given the daily infections at day *t* (DI_t_) and the cumulative infections at day *t* (CI_t_), respectively. The daily growth rate (DGR) was calculated by: DI_t_/CI_t_-1.

Second, the average of the DGR was evaluated. These were determined using the highest values of DI_t_ and CI_t_, weighted for CI_t-1_, and calculated progressively, using the cumulative number of cases from the second day of the series:

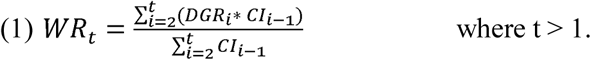

DGR_t_ = DI_t_/CI_t-1_ and DRG_t_ * CI_t-1_ = DI_t_, (1) becomes

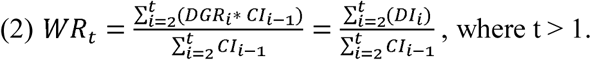

Third, the weighted and cumulated average of the daily growth rate (*WR_t_*) was determined by analyzing the trends in Hubei, from which the results were used to model an epidemic in Italy. The series of *WR_t_* observed in Hubei approach a negative exponential model. Therefore, a nonlinear regression was applied to the exponential decay model (EDM) to evaluate its reliability. In this model, if a function Y(t) is monotonically decreasing with a velocity parameter r>0, then Y(t) the exponential decay model with the form:

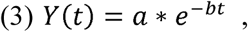

where *t* is time in days, *a* is the starting value Y(0), and *b* the decay rate; in our application, *a* is *WR_t_* when t=0.

To take into account the plateau, which is expected at the end of the epidemic in (3), we added a term *k* that is the rate at the end of the curve:

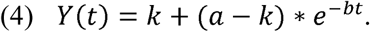

When the number of cases → 0 then zero, *WR_t_* → *k*.

It was thought that the epidemic in Italy had peaked when this model was applied. Therefore, it was assumed that a conceivable *k* term in the exponential decay model for the Italian epidemic could be the same as that observed in the Hubei epidemic that was signed as 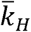, so the model applied was:

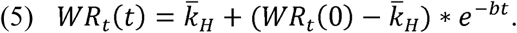

To evaluate the variability of the forecast, we used credibility intervals [7], determined through a first-order Taylor expansion and Monte Carlo simulation [8,9].

Expected occurrences, 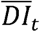, and expected cumulated occurrences 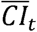, were determined using the inverse of (2) once estimated *WR_t_(t)*.

### Qualitative evaluation of forecast model

A comparison with the Gompertz model, which is often used to estimate cumulative events, was made to evaluate the fitting of the model based on the weighted and averaged growth rates. This was done because the two-parameter Gompertz model (GoM) is based on exponential decay, and is given by the ordinary differential equations:

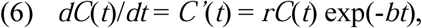

where t is time, C’(t) describes the incidence curve over time, and C(t) is the cumulative number of cases at

time t, and b > 0 describes the exponential decay of the growth rate r (with r > 0). If C(0) is the initial number of cases, then the solution of (6) is

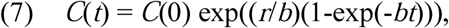

This model was applied to the cumulative infection data in Italy during the same period.

To compare the mathematical models and assess their predictive accuracy at different stages of the outbreak, we estimated the two models (5 and 7) using the observed values in four-time intervals: P1 February 27 to March 17, 2020; P2 February 27 to March 24, 2020; P3 February 27 to March 31, 2020; P4 February 27 to April 07, 2020. We used the estimated parameters at each time period to determine both the rates and the cumulative cases expected, with credibility intervals until May 31, 2020.

It is possible to quantify the error of the model fit to the data using performance metrics [10]. The most widely used performance metrics are the Pearson’s correlation coefficient, root mean squared error (RMSE), mean absolute error (MAE) and mean absolute percentage error (MAPE), given by the respective expressions:

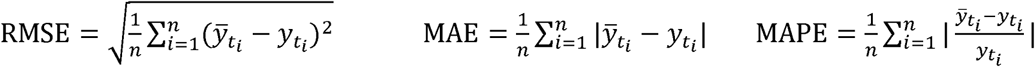

MAPE, together with the correlation coefficient, are performance indicators independent of the outcome size. For this reason, they were used to compare model (5) applied to *WR_t_* model (7) applied to CI. RMSE and MAE were used to compare the same model in relation to the different time phases of the estimate.

These four metrics are also useful for quantifying the error associated with a forecast and were used to evaluate the goodness of fit of the models and to measure the reliability of the model in predicting the values observed after the date of the estimate.

The analysis was carried out using R software (version 3.6.3) [11] with the packages “nls2” [12] and “propagate”[5].

## Results

Starting from February 27, 2020 until April 07, 2020 (last day before the model fitting), the number of infections had grown from 650 to 135,606. The daily growth rate ranged from 2.3% to 62.5%, with a median of 13.4% (Inter Quartile Range IQR 6.9% to 22.8%). The *WR_t_* ranged from 7.0%-45.4%, with a median of 17.3% (IQR, 11.3%-26.0%) (Fig 1).

**Fig 1.**
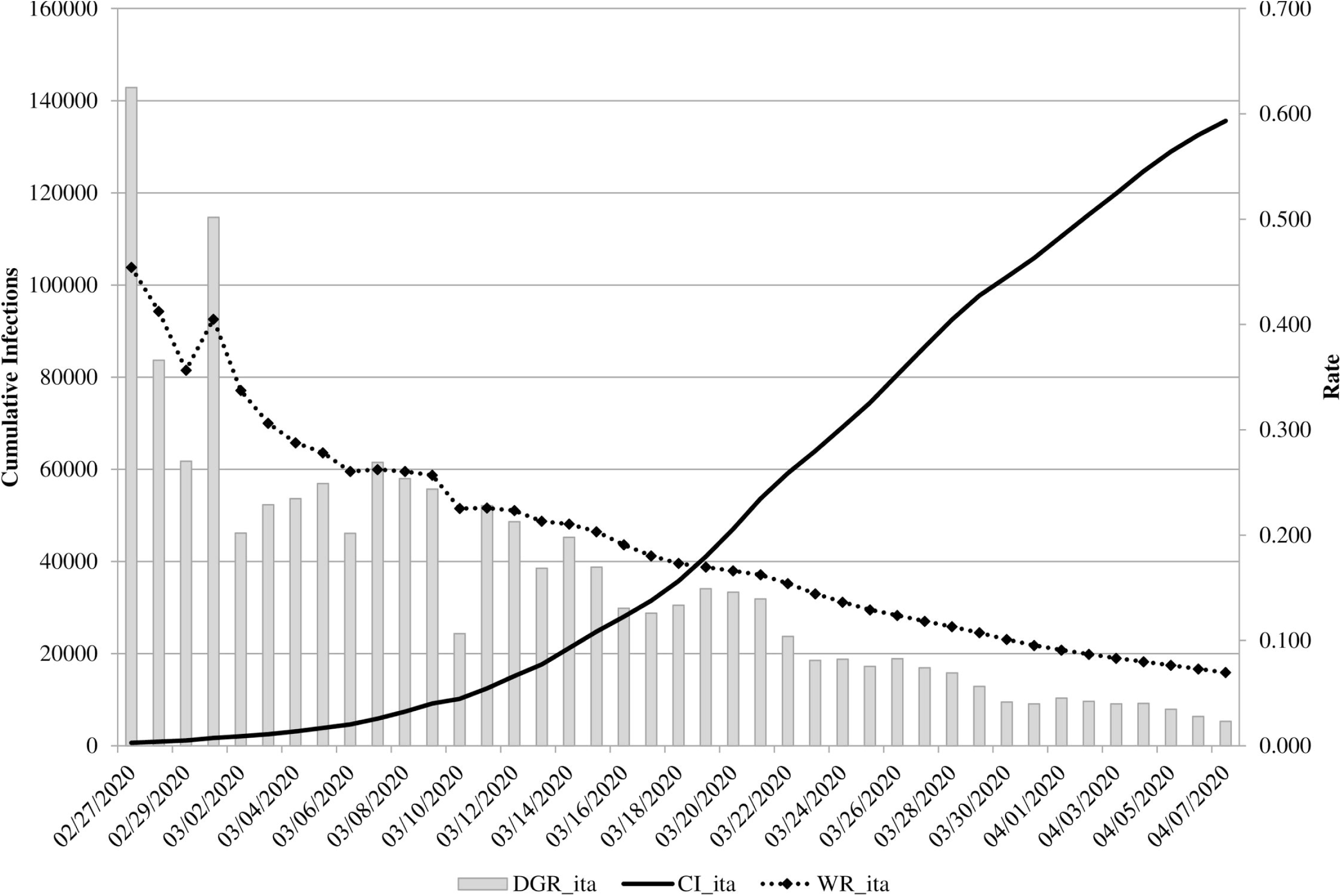
CI, DGR, and *WR_t_* observed and calculated in Italy during the reference period (27 February 2020 - 07 April 2020).

Table 1 shows the cumulative cases, daily growth rates, and *WR_t_* determined at each time point chosen for the evaluated models.

**Table 1.** Descriptive statistics of the CI, DGR, and WR for each time phase

| Time Model<br>(end Date) | N° Days<br>(from 27/02) | CI at end Date<br>(from 650) | DGR (%) | | $WR_t$ (%) | |
| --- | --- | --- | --- | --- | --- | --- |
|  |  |  | Range | Median [IQR] | Range | Median [IQR] |
| P1 (17/03) | 20 | 31,506 | 10.7-62.5 | 22.8 [19.1-25.7] | 18-45.4 | 26.0 [22.1-31.4] |
| P2 (24/03) | 27 | 69,176 | 8.1-62.5 | 20.2 [13.6-24.6] | 13.6-45.4 | 22.5 [17.7-28.3] |
| P3 (31/03) | 34 | 105,792 | 4.0-62.5 | 15.9 [8.8-23.3] | 9.5-45.4 | 20.7 [14.7-26.2] |
| P4 (07/04) | 41 | 135,606 | 2.3-62.5 | 13.4 [6.9-22.8] | 7.0-45.4 | 17.3 [11.3-26.0] |

DGR, daily growth rate; CI, cumulative infections; IQR, inter quartile range; P1–P4, time intervals; $WR_t$ , weighted and cumulated average of the daily growth rate.

To estimate 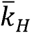 the exponential decay model was applied to the Hubei Province data from January 29, 2020 to March 13, 2020. The initial parameters were k=0, a=0.388, and b=0.05. The algorithm for the estimation of the model converged in three iterations, and all parameters were statistically significant (*a* = 0.406±0.006, p<.0001; *b* = 0.070±0.003, p<.0001; *k* = 0.011±0.004, p=0.019) (Fig 2). The fitting of the curve was remarkably high in relation to the observed values, and the residual standard error was 0.0084.

**Fig 2.**
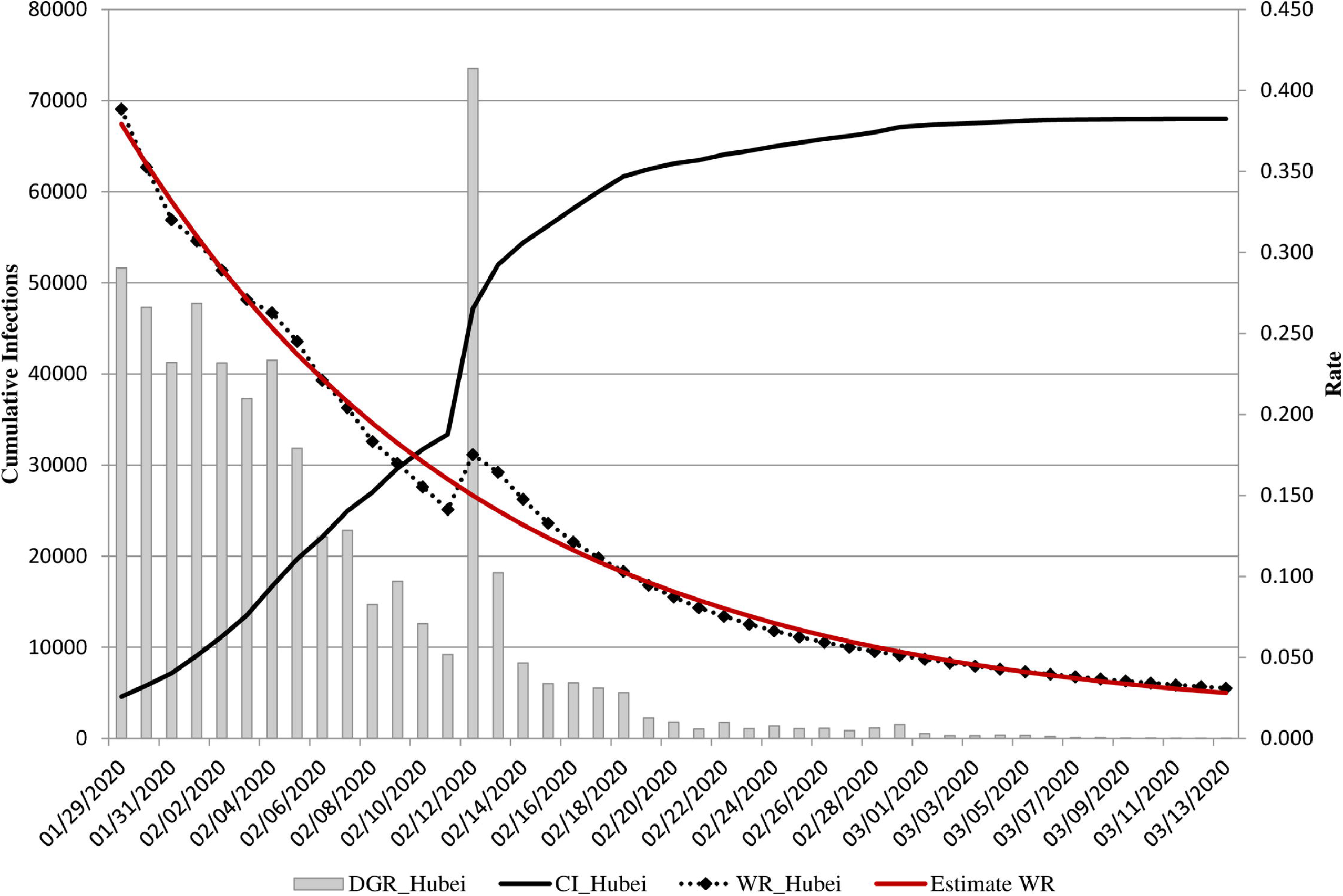
CI, DGR, and *WR_t_* observed and estimated with model 4) in Hubei Province (China) between 29 January 2020 and 13 March 2020.

We used 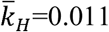 in (5) to estimate *WR_t_* models for the Italian COVID-19 outbreak in the following four-time periods: P1 February 27 to March 13, P2 February 27 to March 17, P3 February 27 to March 31, and P4 March 27 to April 07, 2020. The parameter *a* showed a range from 0.414 to 0.415, which was statistically significant at all time intervals. The parameter *b* ranged between 0.046 and 0.047 and was also statistically significant. The value of RMSE decreased from 0.019 for P1 to 0.014 for P4.

The cumulative number of cases (CI) was estimated at the same time intervals P1-P4 with the model (6): parameters decreased from 602.2 in P1 to 118.2 in P4, whereas exponential decay *b* and growth rate increased (Table 2). In these models, the RMSE increased from 256.7 in P1 to 106 in P2, and the MAE was opposite to the growth rate models (Table 2).

**Table 2.**
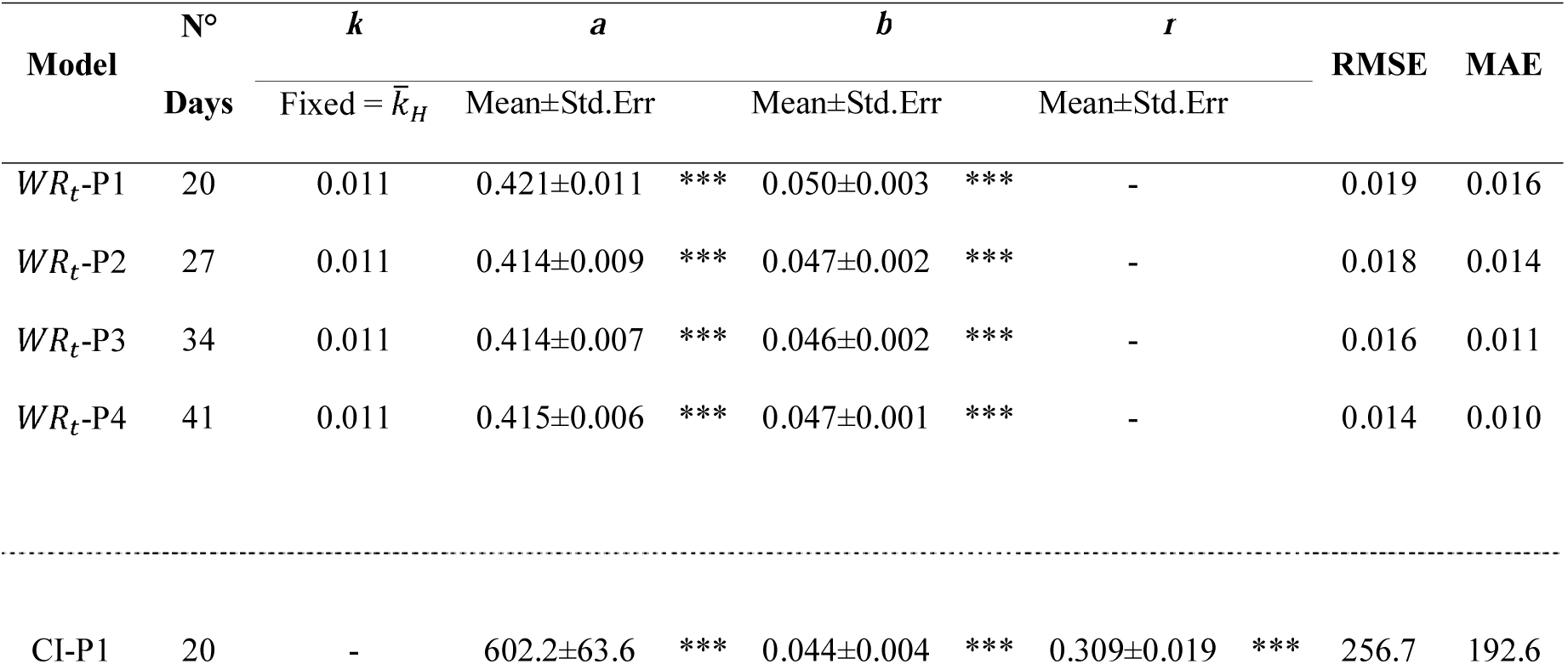
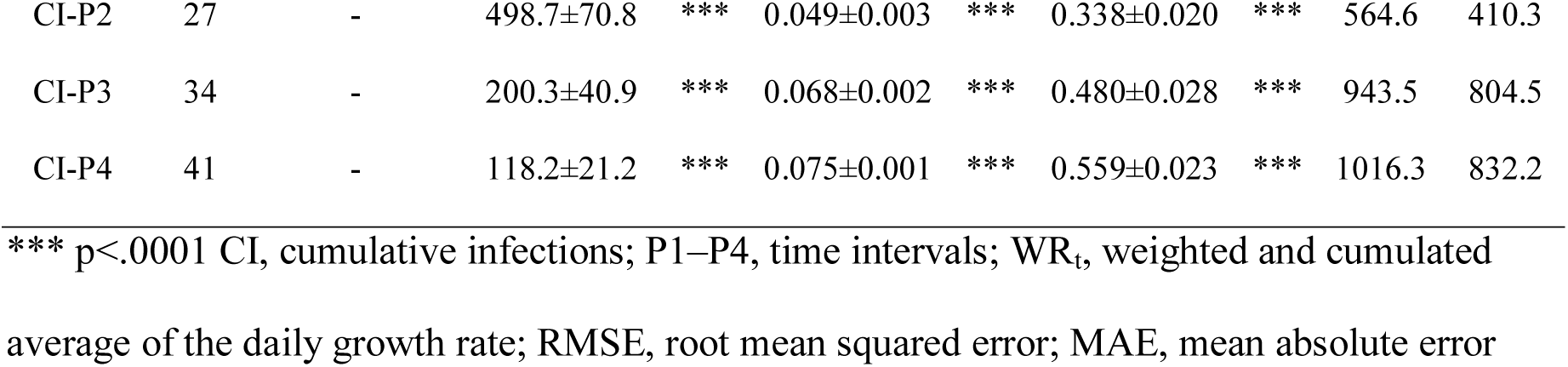
Estimated parameters with the exponential decay model applied to *WR_t_* Eq. (5)) and the Gompertz model applied to the CI (Eq. (7)) for each time phase

The results of the models for *WR_t_* and for CI in the four periods, P1-P4 are shown in Fig 3.

**Fig 3.**
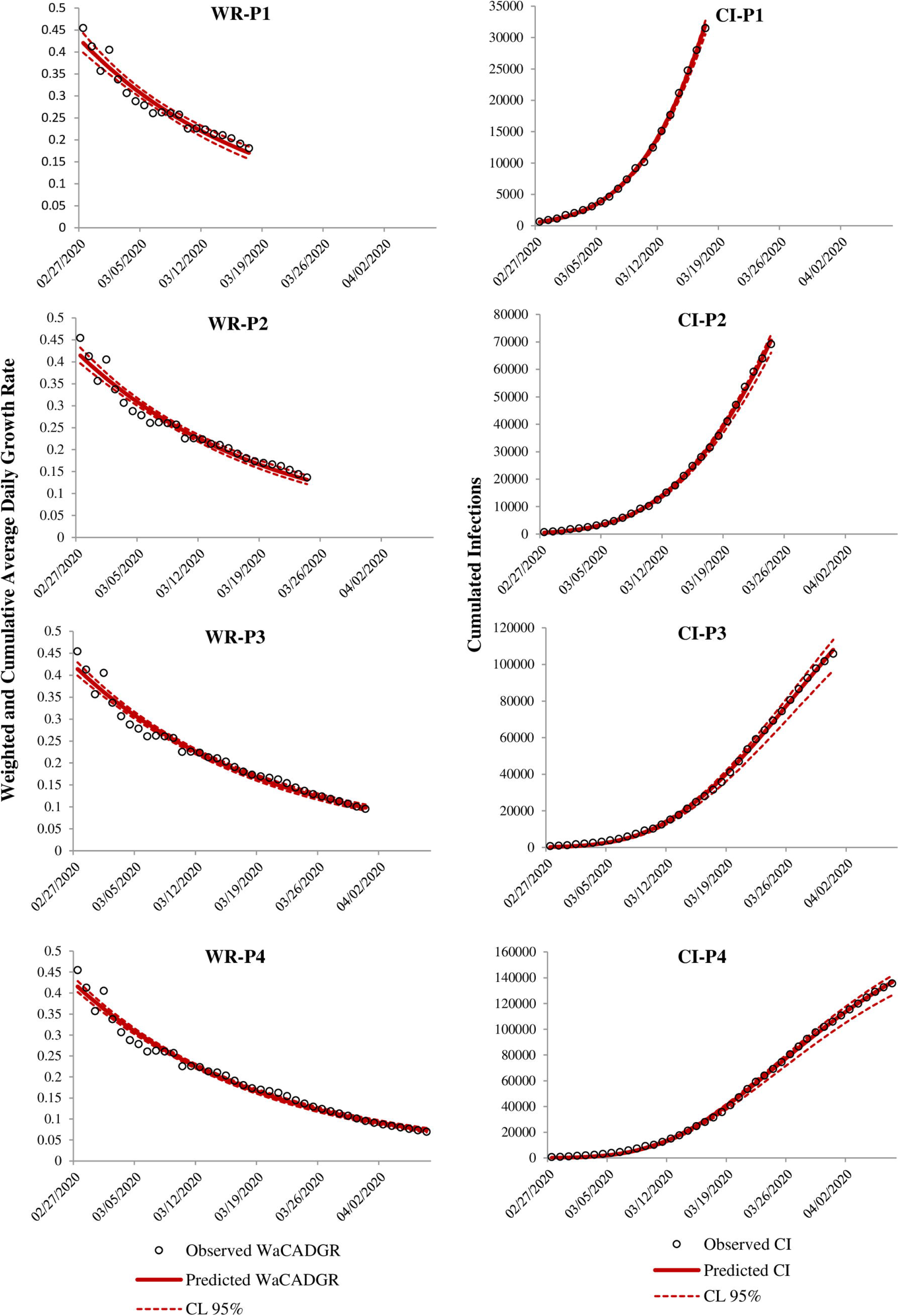
Fitted models based on the exponential decay model (Eq. (5); left column) and Gompertz model (Eq. (7); right column) using epidemic data for the four successive time phases P1, P2, P3, and P4, respectively. The expected values (continuous red line) and the 95% credibility interval (red dotted line) for the fitted models are displayed along with the observed data of the outbreak (empty black circles).

The correlation between the expected and observed values is high in all models, with the CI models having coefficients of nearly 1. The MAPE result was lower only in P1 for the CI models. In contrast, in time intervals P3 and P4, the measure of fit was higher for *WR_t_* than for CI (Table 3 a). Table 3b shows the results of the measure of fit between the forecasted expected values and the observed cases until April 21, 2020. The forecast was made with data already known at intervals P1, P2, P3, and P4, so there were four forecasts made to evaluate what could happen if the models were applied during each period. The MAPE of the exponential decay models applied to *WR_t_* was lower than the Gompertz models for CI, and the correlation coefficient was high for all forecast models. This suggests a strong positive relationship between the expected values forecasted and the observed values.

**Table 3.**
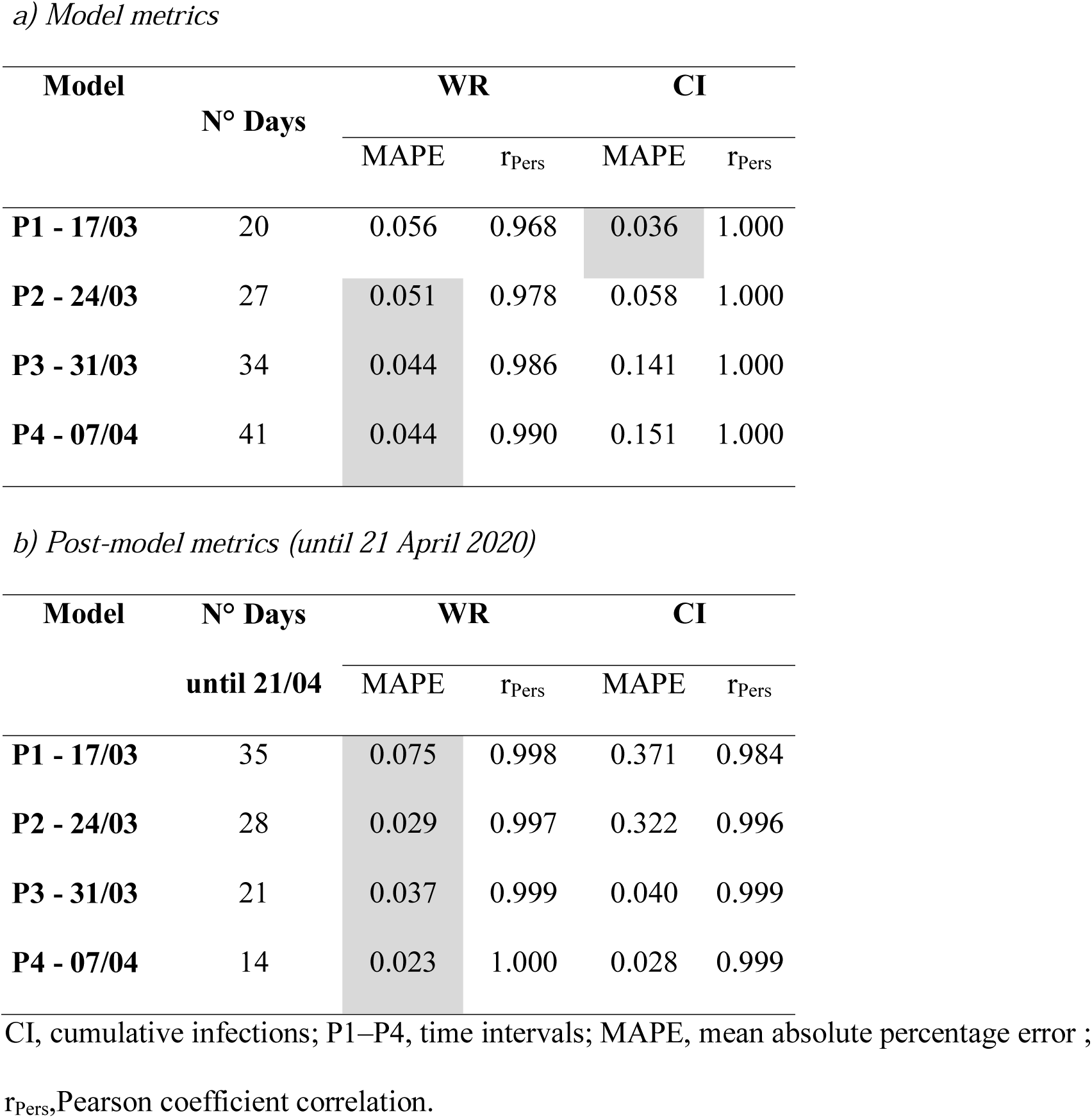
Fitting (a) and forecasting (b) performance statistics for the exponential decay model applied to *WR_t_* (Eq. (5)) and the Gompertz model applied to the CI (Eq. (7)). For each time interval, we highlight the lowest MAPE (gray) for the size of the error.

The forecasts from all models are shown in Fig 4. According to the model *WR_t_* with and the exponential decay with available data for P1, the peak of infections should be reached on April 25 (credibility interval April 12 to May 3) with 131,280 cumulative infections expected (credibility interval 71,415-263,501); with the data available for P2 the peak date should be reached on May 3 (April 23 to May 23) with 197,179 cumulative infections expected (130033-315,269); with data available for P3 the peak should be reached on May 4 (April 25 to May 18) with 202,210 cumulative infections expected (155.235-270,737); with data available for P4 the peak should be reached on May 3 (April 26 to May 11) with 191,586 (160,861-232,023) cumulative infections expected.

**Fig 4.**
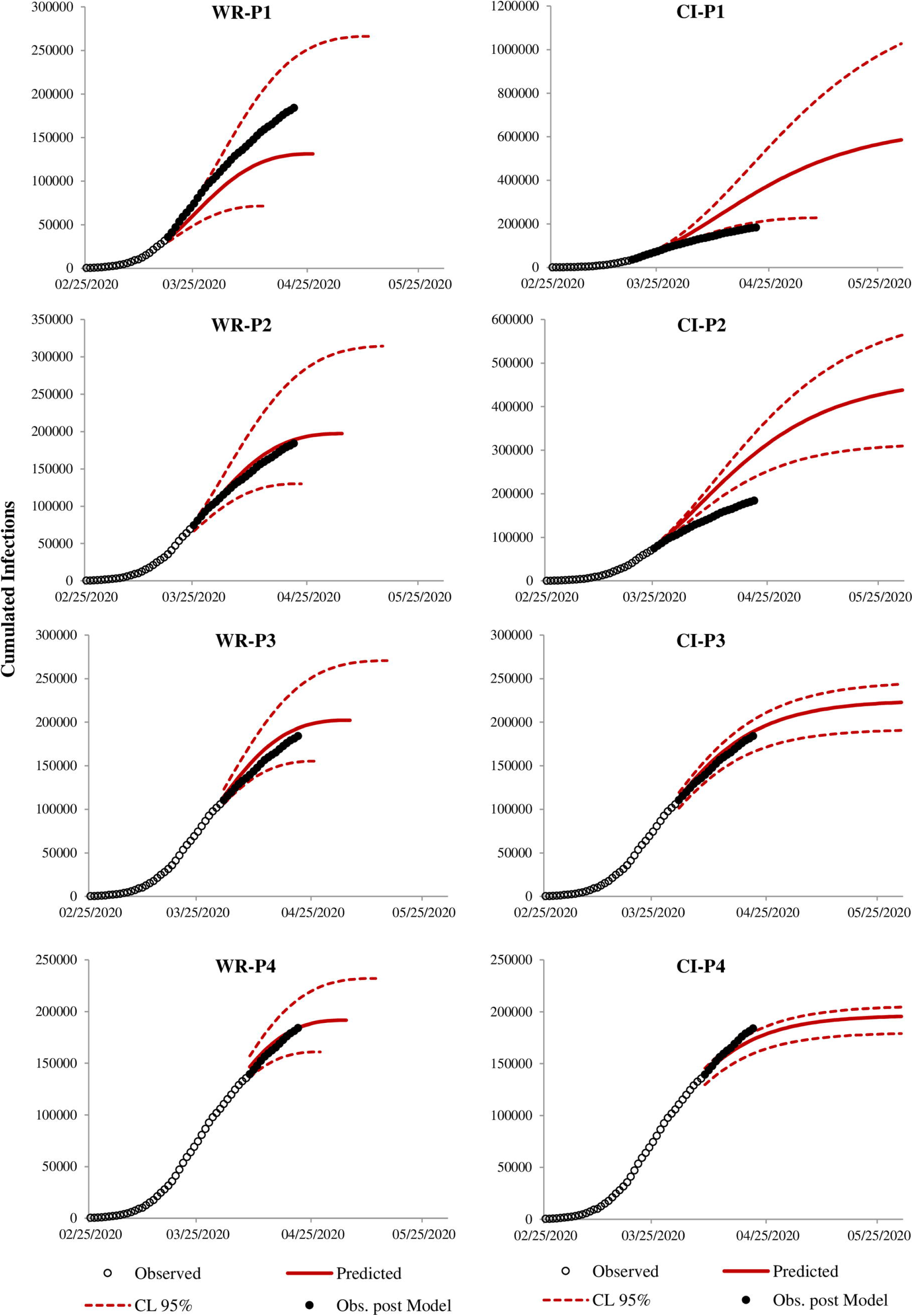
Epidemic forecasts based on the exponential decay model (Eq. (5); left column) and Gompertz model (Eq. (7); right column) calibrated using the epidemic data for scenarios 1, 2, 3, and 4, respectively. The expected values (continuous red line) and the 95% credibility interval (red dotted line) for the calibrated models are displayed along with the observed data of the epidemic, both those used for the estimation of the models (empty black circles) and those after modeling (filled black circles).

All models for CI based on Gompertz have forecast peak dates beyond the May 31 with cumulative infections expected using the data available for P1, P2, P3 and P4 of 585,836 (227,674-1,027,058), 437,665 (309,556-564412), 222,690 (190,579-243,492) and 195,472 (178,952-204,502), respectively.

## Discussion

Dynamic growth models provide an important quantitative framework for predicting epidemic trajectories, generating estimates of key transmission parameters, assessing the impact of control interventions, gaining insight into the contribution of different transmission pathways, and producing short and long-term forecasts [13].

To forecast the evolution of the outbreak, we used phenomenological models with an empirical approach, as such models are useful for reproducing patterns observed in time series data [14]. The result is a reasonably simple temporal description of the epidemic growth patterns [13].

Observation of the epidemic curve in other locales that have already experienced the same outbreak is fundamental in the application of phenomenological models. In this study, we used the epidemic curve of the Chinese province of Hubei to estimate one of the parameters of the exponential decay model that was then applied to the Italian data. It was necessary to find the time point at the beginning of the outbreak in which the curves of the weighted and cumulative average rates were superimposable between the two populations to enable this transposition. *WR_t_* proved to be a stable and useful indicator for estimating the epidemic model based on Chinese data. At the beginning of the epidemic, with few cases, the Gompertz model on the cumulative cases had a better fit and delivered a more precise forecast, supported by the MAPE statistics. However, this does not mean that the forecasted peak date and the number of infections are necessarily correct. The Gompertz model does not have a parameter related to the size of the outbreak. Using data available on 17th of March, the Gompertz model overestimated both the duration and number of cases. The exponential decay model on the same date underestimated the duration and number of cases, but the observed cases were within the credibility limits. Wu et al. showed the same level of inaccuracy using a logistic growth model, generalized logistic growth model, generalized growth model, and generalized Richards model on March 10 [15]. The generalized logistic model provided the best scenario, predicting that the peak would be reached in 10 days. This scenario would have led to a total cumulative number of infected cases of 59,199 (95% CI: [16,796, 107,097]) at 15 days, and 91,719 (95% CI: [16,944, 183,440]) at the end of the pandemic[15]. In contrast, a pessimistic scenario provided by the logistic model predicted that there could be up to 2 million infected cases by the end of May 2020.

The exponential decay model applied to the weighted and averaged growth rates *WR*, seemed to perform better than that shown by Fanelli and Piazza on the 15th of March [16]. They analyzed the temporal dynamics of the COVID-19 outbreak in China, Italy, and France between January 22 and March 15, 2020 and applied a susceptible-infected-recovered-deaths model. They estimated the peak in Italy to be about March 21, 2020, forecasting a maximum number of 26,000 cases at the epidemic peak.

Using data until 24^th^ March, the Gompertz model overestimated the cumulative number of cases. The expected cases in the exponential decay model with *WR* were close to those observed, with a low value of MAPE (0.029).

When the exponential model was applied to data until the 31st of March, the cases forecast was 202,690, whereas the Gompertz model forecast more than 222,690 cases.

Chintalapudi et al. [17] used an ARIMA model, assuming that the lockdown would last for 60 days. According to this study, the 60-day forecast of infected cases could increase to 105,732-182,757. The MAPE for the ARIMA model was 6.25, which is higher compared to the exponential decay model with *WR*.

The exponential model with *WR*, fitted with data available between February 27 and April 07 appears to be close to the observed data, and the MAPE shows a very good fit, with a lower difference between the expected and observed cases than those in the previous model. In contrast, the Gompertz model has a sudden deviation from the observed line and overestimated the number of cases and duration.

It is still unclear what effect the number of swabs performed has on the reported number of positive subjects in the last week of the epidemic compared to at the beginning, and this could significantly affect any modeling of the COVID-19 outbreak.

## Conclusion

An exponential decay model applied to the growth rate of infections weighted with *WR* appears to be useful in estimating the number of cases and peak of the COVID-19 outbreak in Italy.

The model was more reliable in the exponential growth phase due to the statistics used in the model. The epidemic trend information from the COVID-19 outbreak in Hubei Province, China, proved to be essential for estimating the parameters to use in the Italian model.

## Data Availability

All data are fully available without restriction

http://opendatadpc.maps.arcgis.com/apps/opsdashboard/index.html#/b0c68bce2cce478eaac82fe38d4138b1

https://www.who.int/emergencies/diseases/novel-coronavirus-2019/situation-reports

http://www.hubei.gov.cn/zhuanti/2020/gzxxgzbd/zxtb/

## Acknowledgments

We would like to thank Editage (www.editage.com) for English language editing

